# Prevalence and pattern of Post Covid-19 symptoms in recovered patients of Delhi: A Population-Based study

**DOI:** 10.1101/2023.08.25.23294654

**Authors:** Nidhi Bhatnagar, M.M. Singh, Hitakshi Sharma, Suruchi Mishra, Gurmeet Singh, Shivani Rao, Amod Borle, Tanu Anand, Naresh Kumar, Binita Goswami, Sarika Singh, Mahima Kapoor, Sumeet Singla, Bembem Khuraijam, Nita Khurana, Urvi Sharma, Suneela Garg

## Abstract

**Background:** Post-coronavirus disease (COVID) is widely reported but the data of Post COVID-19 after infection with the Omicron variant is limited. This prospective study was conducted to determine the prevalence, pattern, and duration of symptoms related to Covid-19 recovered patients. Methods: Adults (>18 years old) in 11 districts of Delhi who had recovered from Covid-19 were followed up at 3 months and 6 months post-recovery. Results: The study found that the participants had a mean age of 42.07 years, with a standard deviation of 14.89. Additionally, a significant proportion of the participants (79.7%) experienced post-Covid symptoms. The participants elicited a history of Joint Pain (36%), Persistent dry cough (35.7%), anxiousness (28.4%) and shortness of breath (27.1%). The other symptoms reported were persistent fatigue (21.6%), persistent headache (20%), forgetfulness (19.7%) and weakness in limbs (18.6%). The longest duration of symptom was observed in participants reporting anxiousness (138.75 ±54.14) followed by fatigue (137.57±48.33), shortness of breath (131.89±60.21) and joint pain/swelling (131.59±58.76). During the first follow-up, 2.2% of participants had an abnormal ECG reading, while no abnormalities were reported during the second follow-up. Additionally, 4.06% of participants had abnormal chest X-ray findings during the first follow-up, with this number decreasing to 2.16% during the second follow-up. Conclusion: Our study concluded that the clinical symptoms persist in participants until 6 months and a multi-system involvement is seen in the post-COVID period. Thus, the findings necessitate long-term, regular follow-ups.

## 1 Introduction

Severe acute respiratory syndrome 2 coronavirus (SARS-CoV-2), initially diagnosed in China in December 2019, caused a global pandemic. It was associated with an exponential increase in cases and deaths, overwhelming health infrastructure [1]. On 5^th^ May 2023, WHO declared an end to Covid -19 as a Public Health Emergency of International Concern (PHEIC) with >768 million cases and >6.9 million deaths reported to date. [2]. The pandemic triggered a series of epidemiological studies to understand disease transmission, prevention and control measures. The variability of disease based on strain in circulation and poor understanding of pathophysiology resulted in many trial treatments during the phase of outbreak surge. The wide clinical spectrum of SARS-CoV-2 also made the disease complex to understand due to multi-organ involvement [3] [4].

The acute Covid-19 infection across the literature is reported to last up to 4 weeks before recovery [5]. However, there is growing evidence that signs and symptoms related to Covid-19 infection continue to persist post-recovery in a significant proportion of cases. The condition characterized by the continued existence of symptoms after the initial infection and recovery from Covid-19 has been termed “Long-Covid”, “Post-covid” and “Post-acute Covid-19 syndrome” [5]. There is a lack of consensus on the characteristics defining this condition. The British Medical Association defines a syndrome “as a set of medical signs and symptoms which are correlated with each other and associated with a particular disease’’ [6] The National Institute of Health and Care Excellence (UK) defined post-covid-syndrome as “a cluster of signs and symptoms that develop during or after an infection consistent with COVID-19, continue for more than 12 weeks and are not explained by an alternative diagnosis.” [5] The long-term symptoms of Covid-19 can involve cardiopulmonary symptoms (decline in respiratory function, fibrosis, pleural involvement, myocarditis, pericardial effusion), post covid-19 thrombosis, immune-mediated manifestations (arthritis, myositis, pancreatitis), and other skin, neurological, renal, haematological, endocrine and systemic manifestations [5, 7]. The impact of persistent Covid-19 symptoms is reported significant on the mental health and emotional well-being of patients [8].

To date, there has been limited exploratory research on post-covid syndrome with scarce data on long-term outcomes among recovered Covid-19 patients of emerging economies. [8] [9]. We, therefore, planned to conduct this study to assess post-covid phenomenon or symptomatology in recovered Covid-19 patients in Delhi, India. The objective of this research was to determine the prevalence, pattern, and duration of symptoms related to Covid-19 recovered patients.

## 2 Methodology

### Study Design

A Prospective study was conducted in 11 districts of Delhi among recovered Covid-19 patients. The study subjects were enrolled after recovery and followed up at 3 months and 6 months post-recovery.

Operational definitions used in the study

1. Recovery (Hospitalized Patients): defined as Covid patients discharged from the hospital after treatment completion.
2. Recovery (Non-Hospitalized Patients) is defined as a duration of 14 days from the symptom onset since the confirmation of diagnosis using RT-PCR or antigen tests.
3. Acute COVID-19: Symptoms up to 4 weeks since diagnosis of SARS-CoV-2[5]
4. Ongoing symptomatic COVID: Symptoms from 4 to 12 weeks since diagnosis of SARS-CoV-2. [5]
5. Post-COVID: Symptoms developed during or after infection and continue for more than 12 weeks.[5]
6. Long COVID: Signs and Symptoms that continue for more than four weeks (Ongoing symptomatic COVID and post-COVID syndrome subgroups) [5]

Study area and period

The study was carried out from October 2021 to July 2023 (22 months) in 11 districts of Delhi, India.

Inclusion criteria

1. Adult individuals (>18 years) with a history of mild, moderate and severe Covid-19 after 14 days of diagnosis.
2. Adult individuals with a history of Covid infection within the last 6 months.

Exclusion criteria

1. Pregnant women
2. History of re-infection with Covid-19
3. Patient in the terminal stage of any disease.

### Sample size and sampling technique

The reported prevalence of long COVID syndrome is 22%. Considering the prevalence of long COVID syndrome at 22%, 95% confidence levels, and relative precision as 4.4%, and considering 20% allowable error and 20% loss to follow-up, the sample size was estimated at 410 [10]. By the Per-Protocol principle, the analysis of the collected data is based on the second follow-up sample size (369).

### Data collection procedure

The study participants were diagnosed with Covid-19 in the period ranging from April 2022 to September 2022. A daily list of Covid-19 recovered patients that were hospitalized and those managed through home isolation during the course of their illness was obtained from the Directorate General of Health Services, Government of NCT, Delhi. The data was cleaned by removing individuals with missing addresses, contact numbers and aged less than 18 years. Thereafter, the participants who met the inclusion criteria were contacted through phone calls and consequently household visits were done. They were enrolled in the study after obtaining written and informed consent for prospective follow-up at baseline, 3- and 6-months post-recovery. A detailed medical history and information on the past episode of Covid-19 were obtained through the participant interview. A comprehensive history taking was done for all the participants at each visit to their households. For baseline assessment, socio-demographic characteristics of the participants were collected using an interview schedule including age, sex, education, socio-economic status, healthcare-seeking behaviour for COVID episode, baseline comorbidity details and treatment profile at their household. Furthermore, the standardized and validated WHO case report form for post-covid conditions was exercised for participant information.[11] The multidisciplinary team of doctors reviewed the medical history and conducted investigations and clinical assessments of study participants to provide advice on further biochemical investigations, referrals, and treatment. The blood samples were tested in a government-approved laboratory with participant consent using the Vitros 5600 integrated clinical chemistry and immunoassay analyzer, and the Stago STA MAX coagulation analyzer. [Annexure]

### Statistical analysis

The data was entered in MS Excel and analyzed in IBM SPSS Version 25. Results were expressed in frequency and proportions for categorical variables, median (IQR) if non-normal continuous data, and mean (SD) if continuous normal data. Prevalence and incidence of post-covid related symptoms and consequences were assessed during the baseline and each follow-up visit. A per-protocol analysis was followed as there is less attrition in the follow-up period. Standard cut-off scores were applied to assess for the presence of depression, anxiety, poor sleep quality, and suboptimal quality of life which comprised the dependent variables. The independent variables included age, sex, severity of Covid-19, duration of hospitalization, and history of tobacco smoking and substance abuse. Comparison to assess the difference in means between two groups was assessed using students’ unpaired t-test or the Mann-Whitney U test and more than two groups assessment at 3 points was done by repeated measures ANOVA (Tukey’s post hoc test) or Freidman test as per the data distribution. A p-value < 0.05 was considered statistically significant.

## 3 Results

The study was conducted among 413 participants. The response rate was 379(91.76%) at the first follow-up and 369(89.34%) at the second follow-up. By the Per-Protocol principle, the analysis of the collected data is based on the second follow-up sample size (369). The mean age of the study participants is 42.07(14.89) years. The socio-demographic profile of the subjects concerning gender, educational status, history of smoking and alcohol consumption was studied. Nearly half (50.13%) of the participants were male, (97.84%) were literate, (92.96%) had never smoked and (9.21%) had a history of alcohol consumption. The majority of the (93.5%) participants were vaccinated and 26.56% had any co-morbidity (Table 1). There was a significant association of gender and age with the post-covid syndrome (<0.05).

**Table 1:**
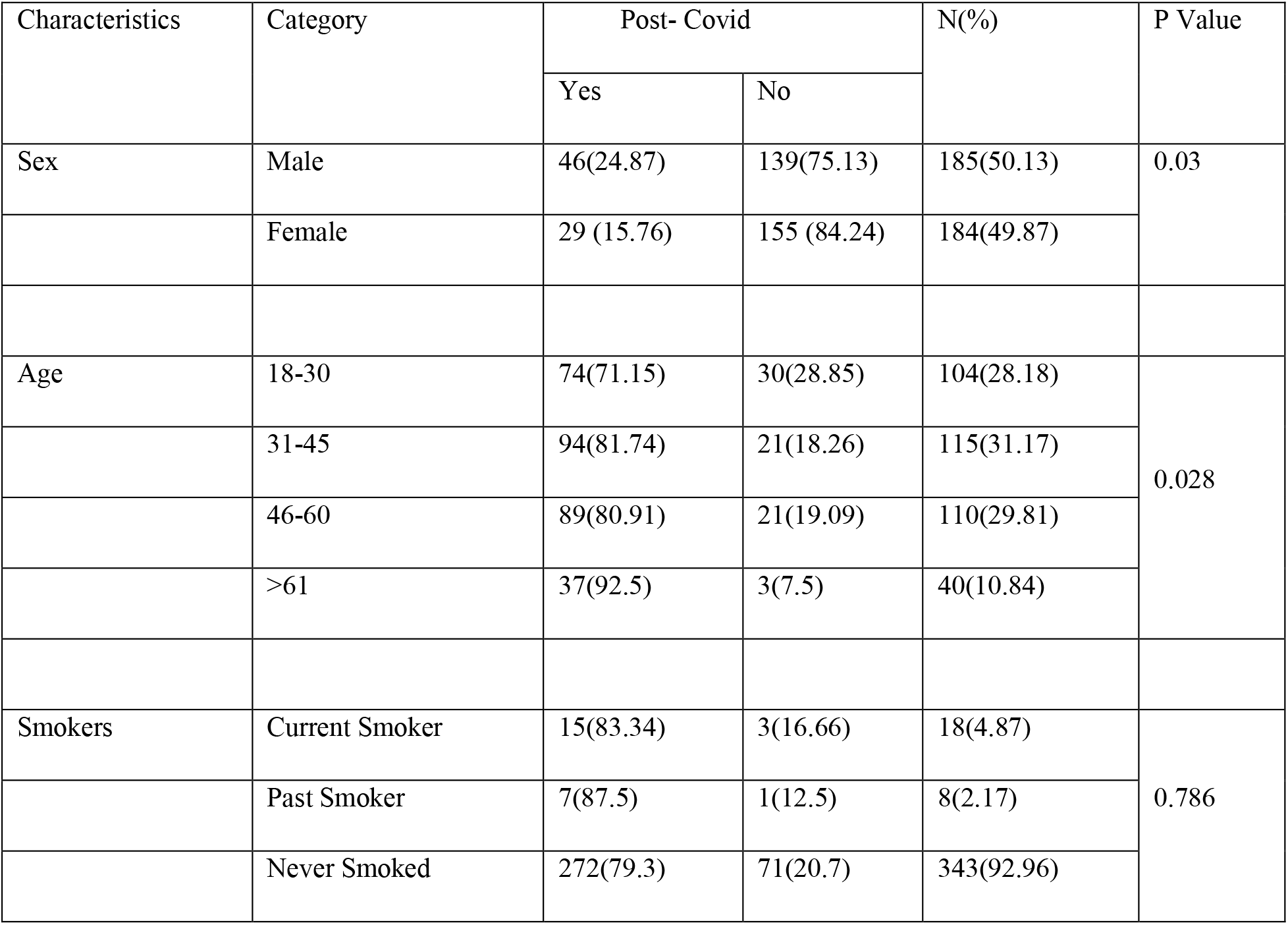
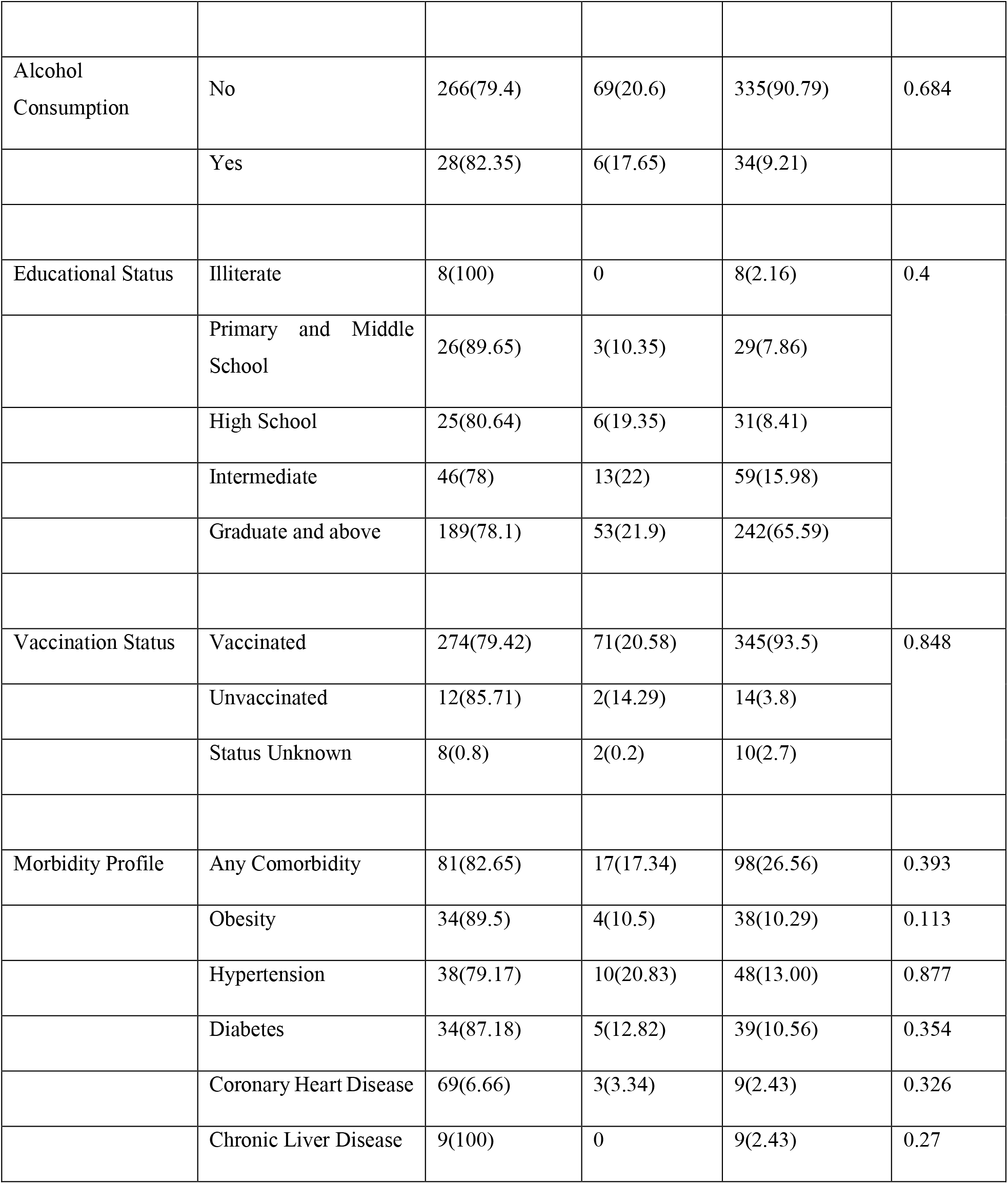
Socio-Demographic Profile of recovered Covid-19 patients. (N=369)

| Characteristics | Category | Post- Covid |  | N(%) | P Value |
| --- | --- | --- | --- | --- | --- |
|  |  | Yes | No |  |  |
| Sex | Male | 46(24.87) | 139(75.13) | 185(50.13) | 0.03 |
|  | Female | 29 (15.76) | 155 (84.24) | 184(49.87) |  |
| Age | 18-30 | 74(71.15) | 30(28.85) | 104(28.18) | 0.028 |
|  | 31-45 | 94(81.74) | 21(18.26) | 115(31.17) |  |
|  | 46-60 | 89(80.91) | 21(19.09) | 110(29.81) |  |
|  | >61 | 37(92.5) | 3(7.5) | 40(10.84) |  |
| Smokers | Current Smoker | 15(83.34) | 3(16.66) | 18(4.87) | 0.786 |
|  | Past Smoker | 7(87.5) | 1(12.5) | 8(2.17) |  |
|  | Never Smoked | 272(79.3) | 71(20.7) | 343(92.96) |  |
| Alcohol Consumption | No | 266(79.4) | 69(20.6) | 335(90.79) | 0.684 |
|  | Yes | 28(82.35) | 6(17.65) | 34(9.21) |  |
| Educational Status | Illiterate | 8(100) | 0 | 8(2.16) | 0.4 |
|  | Primary and Middle School | 26(89.65) | 3(10.35) | 29(7.86) |  |
|  | High School | 25(80.64) | 6(19.35) | 31(8.41) |  |
|  | Intermediate | 46(78) | 13(22) | 59(15.98) |  |
|  | Graduate and above | 189(78.1) | 53(21.9) | 242(65.59) |  |
| Vaccination Status | Vaccinated | 274(79.42) | 71(20.58) | 345(93.5) | 0.848 |
|  | Unvaccinated | 12(85.71) | 2(14.29) | 14(3.8) |  |
|  | Status Unknown | 8(0.8) | 2(0.2) | 10(2.7) |  |
| Morbidity Profile | Any Comorbidity | 81(82.65) | 17(17.34) | 98(26.56) | 0.393 |
|  | Obesity | 34(89.5) | 4(10.5) | 38(10.29) | 0.113 |
|  | Hypertension | 38(79.17) | 10(20.83) | 48(13.00) | 0.877 |
|  | Diabetes | 34(87.18) | 5(12.82) | 39(10.56) | 0.354 |
|  | Coronary Heart Disease | 69(6.66) | 3(3.34) | 9(2.43) | 0.326 |
|  | Chronic Liver Disease | 9(100) | 0 | 9(2.43) | 0.27 |

Nearly half of the participants (47.5%) had Long Covid symptoms and (79.7%) had post-Covid symptoms. (Figure 1) The participants had a history of Joint Pain (36%), Persistent dry cough (35.7%), anxiousness (28.4%) and shortness of breath (27.1%). The other symptoms reported were persistent fatigue (21.6%), persistent headache (20%), forgetfulness (19.7%) and weakness in limbs (18.6%). (Table 2) The longest duration of symptom was observed in participants reporting anxiousness (138.75 ±54.14) followed by fatigue (137.57±48.33), shortness of breath (131.89±60.21) and joint pain/swelling (131.59±58.76). A prolonged duration of symptoms was also observed in patients with body pain (127.48±47.44), dry cough (121.01±56.48) and weakness in limbs (120.91±49.34). Anxiousness and persistent dry cough were significantly found to be of longer duration in males (p<0.05). Weakness in limbs and fever were observed to be of significantly longer duration among the vaccinated (p<0.05). The other reported symptoms reported were forgetfulness (116.47±52.86) and headache (114.74±42.68). The forgetfulness was seen to be of significantly longer duration among participants reporting any co-morbidity (Table 3).

**Table 2:** Distribution of study participants on the basis of prevalence of post-Covid symptoms (N=369)

|  | Baseline<br>(N=413) | Follow up 1 (N=379) |  | Follow up 2 (N=369) |  | PCoVS<br>Prevalence |
| --- | --- | --- | --- | --- | --- | --- |
| Symptoms |  | Prevalence | Incidence | Prevalence | Incidence |  |
| Any Symptom | 196(47.5%) | 228 (61.8) | 107 (29.0) | 229 (62.1) | 39 (10.6) | 294 (79.7) |
| Anxiousness | 41(9.9%) | 84 (22.8) | 70 (19.0) | 43 (11.7) | 14 (3.8) | 105 (28.4) |
| Join Pain/ Swelling | 40(9.7%) | 75 (20.3) | 67 (18.2) | 89 (24.1) | 47 (12.7) | 133 (36.0) |
| Persistent Fatigue | 67(7.2%) | 66 (17.9) | 44 (11.9) | 26 (7.0) | 11 (3.0) | 80 (21.6) |
| Shortness of breath | 28(6.8%) | 52 (14.1) | 38 (10.3) | 74 (20.1) | 45 (12.2) | 100 (27.1) |
| Persistent body pain | 46(11.1%) | 46 (12.5) | 40 (10.8) | 48 (13.0) | 31 (8.4) | 83 (22.4) |
| Weakness in limbs | 40(9.7%) | 48 (13.0) | 40 (10.8) | 34 (9.2) | 17 (4.6) | 69 (18.6) |
| Forgetfulness | 17(4.6%) | 38 (10.3) | 34 (9.2) | 44 (11.9) | 30 (8.1) | 73 (19.7) |
| Persistent dry cough | 46(11.1%) | 38 (10.3) | 30 (8.1) | 112 (30.4) | 84 (22.8) | 132 (35.7) |
| Persistent headache | 27(7.31%) | 37 (10.0) | 32 (8.7) | 45 (12.2) | 33 (8.9) | 74 (20.0) |

**Table 3:**
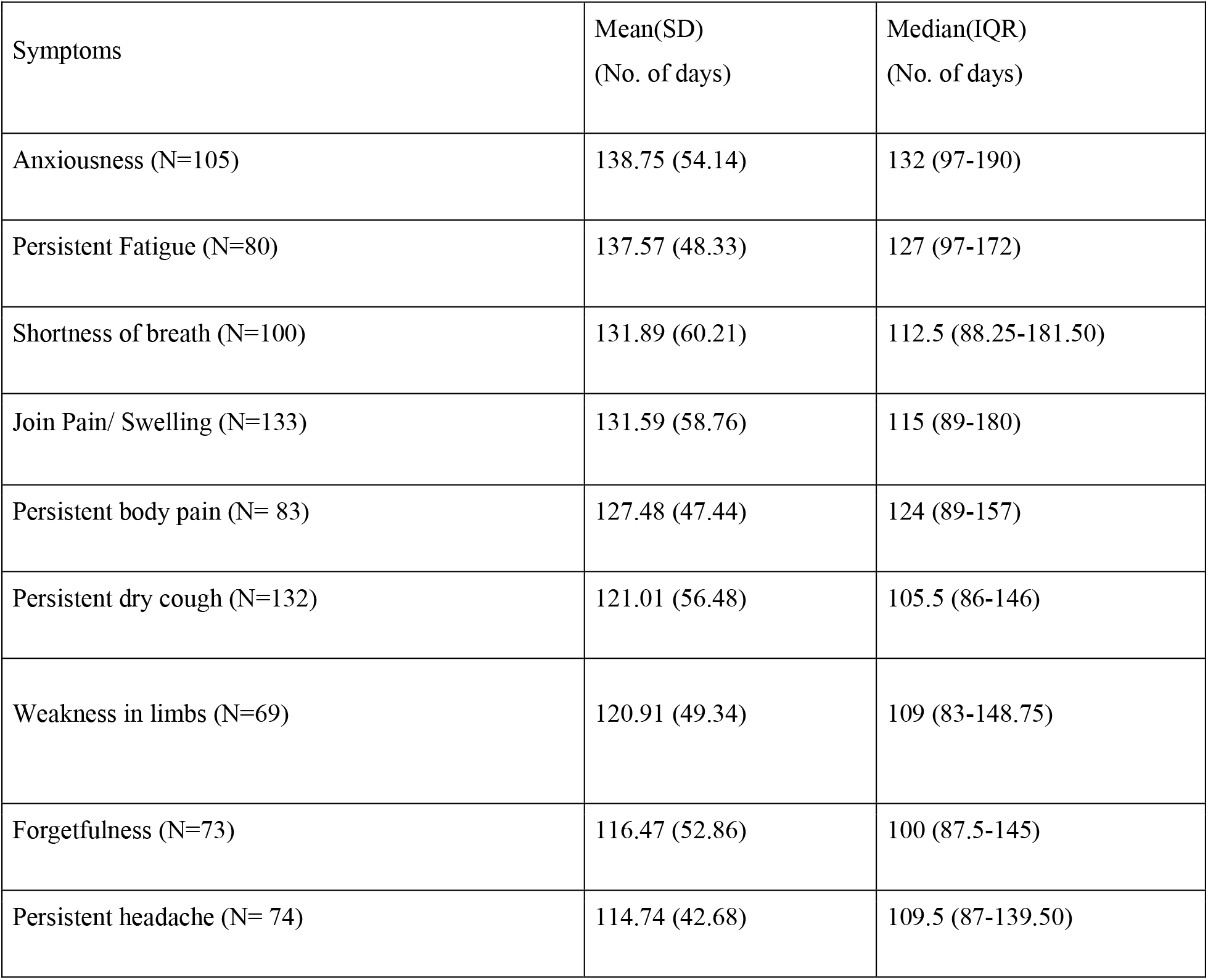
Duration of post-COVID symptoms in study participants.

**Figure 1.**
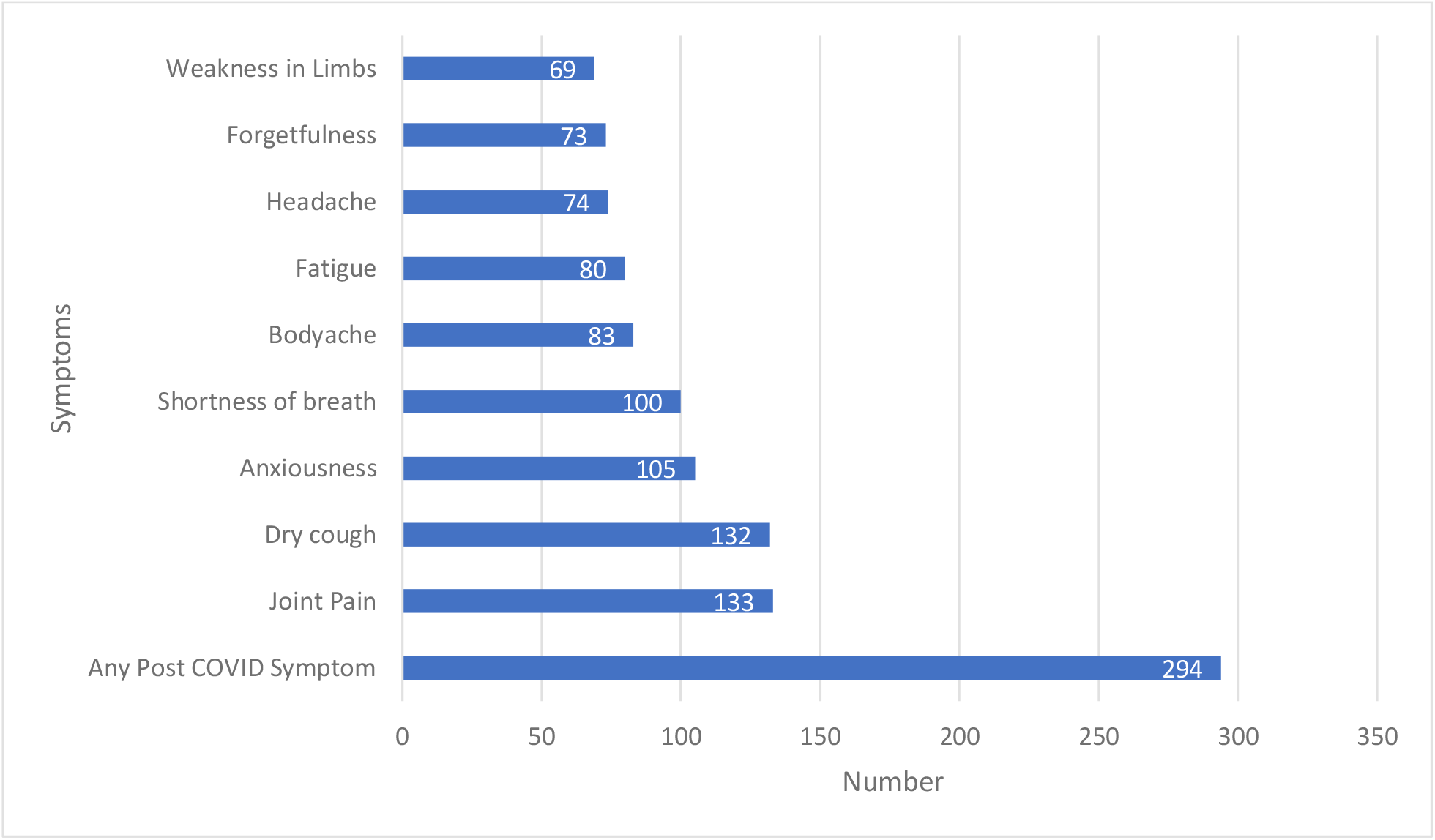
Pattern of post-Covid symptoms (N=369)

The biochemical investigations were done for study participants at baseline, first follow-up and second follow-up. The Shapiro-Wilk test of normality presented the data to be skewed in the distribution of the biochemical parameters. Based on the data analyzed, there does not appear to be a significant difference in the median levels of TSH, HbA1c, AST/SGOT, ALT/SGPT, ALP, Creatinine, urea, and uric acid between individuals with Post Covid syndrome and those without. (p>0.05). The median values for all parameters were within the normal range and were comparable between the two groups. [Table 4] The new cases of raised HbA1C were detected in (3.5%) at the first follow-up and (6.9%) at the second follow-up. New cases of elevated TSH were observed in (9.2%) at the first follow-up and (6.6%) at the second follow-up. In the liver function tests, AST/SGOT was raised in (4.2%) of the participants at the first follow-up and (3.6%) at the second follow-up. The ALT/SGPT was elevated in (8.4%) of participants at the first follow-up and (4.8%) at the second follow-up. Furthermore, at the first follow-up, ALP was detected elevated in (12.6%) and at the second follow-up it was raised in (7.0%). Under the Kidney function test panel, the patients were investigated for Creatinine, urea and uric acid. Creatinine level was raised in (0.8%) at the first follow-up and (1.1%) at the second follow-up. Only (2.5%) reported elevated urea at the first follow-up and (1.6%) at the second follow-up. Likewise, just (1.9%) reported high uric acid levels at follow-up 1 and (1.4%) at follow-up 2.

**Table 4:** Association between biochemical parameters and Post Covid syndrome.

| Biochemical Parameters | First Follow-Up |  |  | Second Follow-Up |  |  |
| --- | --- | --- | --- | --- | --- | --- |
|  | Post-Covid Symptoms |  | 'P Value'* | Post-Covid Symptoms |  | 'P Value'* |
|  | Present | Absent |  | Present | Absent |  |
| HbA1C(> 6.5%) | 5.4(0.9) | 5.3(0.6) | 0.334 | 5.35(1.2) | 5.2(0.4) | 0.241 |
| TSH(>4.68mIU/L) | 2.98(2.16) | 2.78(2.01) | 0.385 | 3.07(1.83) | 2.99(2.11) | 0.284 |
| AST (>59 IU/L) | 33(12) | 33(14) | 0.943 | 33(14) | 34(15) | 0.993 |
| ALP(>126 IU/L) | 95(34) | 100(40) | 0.235 | 98(36) | 101.5(34) | 0.298 |
| ALT(>50 IU/L) | 30(22) | 29(20) | 0.420 | 32(21) | 30(21) | 0.364 |
| Creatinine<br>(>1.3mg/ dl) | 0.7(0.3) | 0.8(0.2) | 0.103 | 0.80(0.2) | 0.80(0.2) | 0.233 |
| Urea(>43 mg/dl) | 23(10) | 25(9) | 0.428 | 26(7) | 25(5) | 0.177 |
| Uric Acid(8.5mg/dl) | 5.3(2.15) | 5.35(2.2) | 0.710 | 5.45(2.0) | 5.1(2) | 0.518 |

\*Man Whitney U Test

The peripheral blood smear test was done for the patients at baseline, first and second follow-up. The presence of transformed lymphocytes and activated Monocytes was seen in (1.1%) of the participants at the first follow-up 1 and (0.8%) in the second follow-up. The Absolute Neutrophil Count was seen in the peripheral smear of (16.5%) at the first follow-up and (11.7%) at the second follow-up. The Absolute Eosinophil Count was reported in (7.9%) at the first follow-up and (4.6%) at the second follow-up. The ECG was reported as abnormal in (2.2%) of the participants at the first follow-up where blocks (37.5%), bradycardia (25%), low voltage complex (12.5%), Q wave (12.5%) and arrhythmia (12.5%) formed the diagnosis. No abnormality in ECG was reported at the second follow-up. At the first follow-up, (4.06%) of participants had abnormal chest X-ray findings. These findings were reported in the parenchymal (66.67%), pleural (26.67%) and mediastinal (6.66%) regions. At the second follow-up, (2.16%) had abnormal findings showing parenchymal (62.5%), pleural (12.5%) and mediastinal (25%) region involvement (Figure 2).

**Figure 2.**
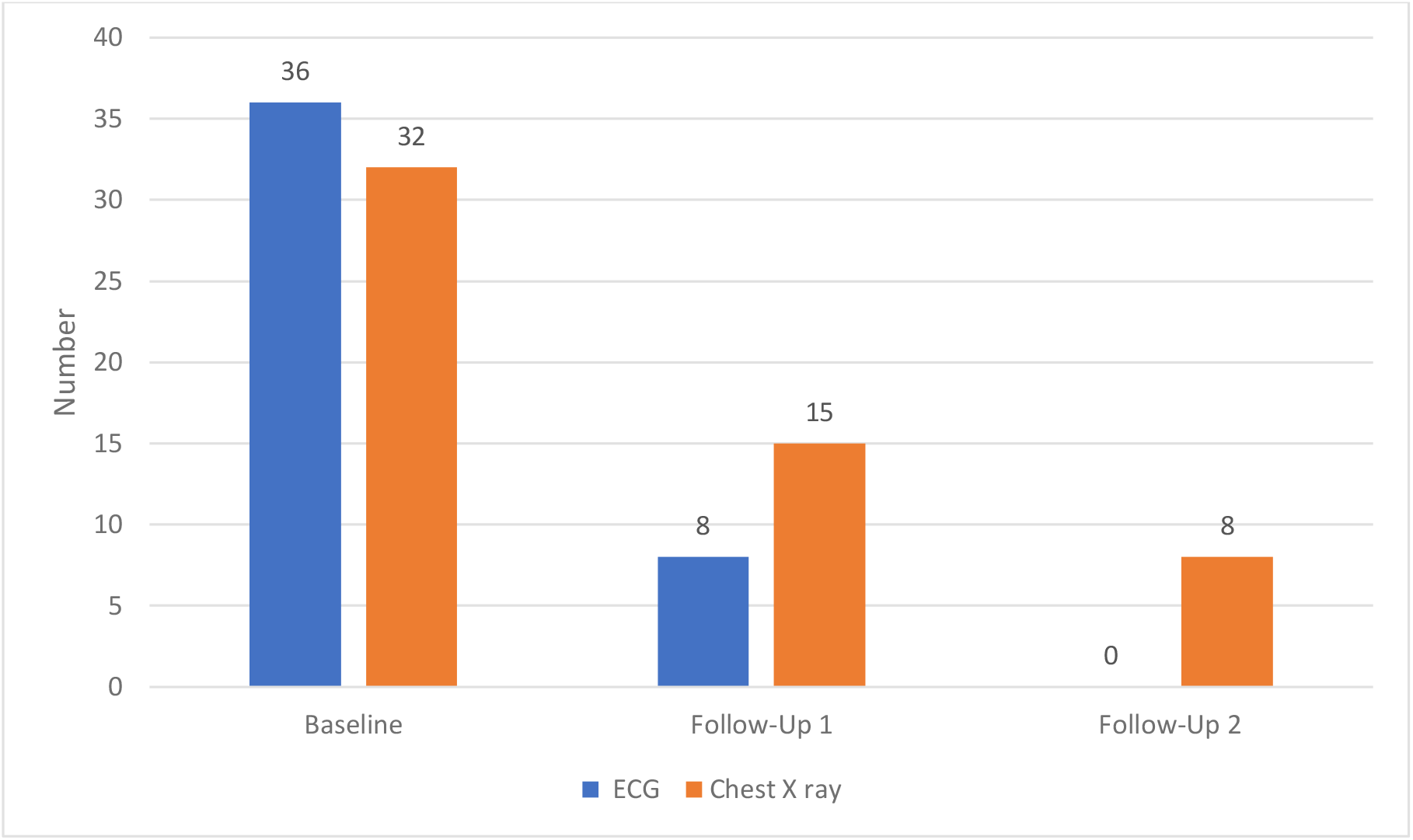
Number of participants with ECG and Chest X-Ray abnormalities. (N=369)

## 4 Discussion

Our study provides insight into the symptomology which could develop in the long term in the Covid-19 diagnosed patients. To the best of our knowledge, ours represents one of the first community-based prospective cohort study to assess post-covid syndrome in India. In our cohort of participants, the prevalence of post covid syndrome was alarming with nearly two-third of the participants reporting one or more post covid symptoms. A similar finding was observed in a study conducted by Huang C in China in 2020 with nearly two-thirds (68%) of the participants reporting at least one post-covid 19 symptom.[12] Another prospective cohort study conducted by Sevalkumar J in 2020 in Norway observed nearly half (48.5%) of the participants with a post-covid syndrome which is lower than our estimate.[13] This could be due to the difference in age profile of the participants observed in the study which was 12-25 years and their study was conducted in Norway population with B.1.1.7 (Alpha) variant of SARS-CoV-2 dominant in the geographical area in 2020 whereas in our study Omicron variant of SARS-CoV-2 was predominant during the study period. Few other studies conducted globally with a similar objective found a higher prevalence of Post covid 19 in participants. [14, 15][Table 1]

In the study, (25.2%) of participants were diagnosed with one or more co-morbidity at baseline and (26.56%) at follow-up 2. A similar finding is observed in a study conducted by Menges D et al (2021) with (34%) of participants reporting at least one chronic comorbidity at baseline [16]. Our study reported (12.1%) with Hypertension and (9.7%) with Diabetes Mellitus. A similar study conducted by Cioboata R (2020) found (36%) of the participants with Hypertension and (14.52%) with Diabetes.[17] The difference could be due to alpha strain being reported in the latter study period. In the current study, (93.5%) of the participants were vaccinated (2 doses) at the time of baseline data collection. The finding is similar to the reported national data coverage of 90% (2022). There was no statistically significant difference found between the vaccination and unvaccinated cohort related to post-covid symptoms. [Table 1]

In our study, we found that there was no statistically significant difference in the LFT and KFT parameters between the groups of individuals who had Post Covid Syndrome and those who did not. The reported values were found to be comparable between the groups. Similar findings were observed in the study conducted by Alfadda A et al in 2019 to investigate the clinical and biochemical parameters in people who recovered from Covid 19 post 6 months. [18] In the current study, we found the lymphocyte counts to decrease from baseline to the second follow-up. Similar findings were observed in a study conducted by Gameil et al in 2021 to assess long-term biochemical residue after covid-19 recovery.[19] Similarly, in our study monocytes also declined in the participants with follow-up. A similar finding is seen in a study conducted by Ruenjaiman V et al to study the response of innate immune cells post-recovery from SARS-CoV-2 infection. [20] We found (4.06%) of participants with abnormal chest X-ray findings at the 3 months follow-up period. A study conducted by Fogante, M et al reported (50.4%) of the subjects reported chest X-rays abnormalities at 3 months follow-up which is higher than our reported result.[21] It’s possible that the differences in the findings are due to the methodology used. Specifically, the latter study focused on hospitalized patients may have produced different results than our study. In our study, only 2.2% of the participants reported ECG changes at the first follow-up and no changes in ECG were found at the second follow-up. A study was conducted by Øvrebotten et al. to assess the changes in cardiac structure and function 3-12 months after hospitalization for Covid-19. Regardless of the severity and persistent dyspnea, cardiac structure and function remained unchanged. [22]

There are a few limitations of our study. The non-availability of baseline clinical profiles of the study participants before Covid-19 infection was a major limitation. Second, as the participants have self-reported the symptoms experienced post covid, recall bias is possible. Third, in this study, we could not distinguish whether the post-covid symptoms were due to covid vaccination, re-infection of covid, or post-Covid syndrome. Fourth, even though we are assuming the omicron strain to be prevalent during the study period, we have no conclusive evidence for it.

## 5 Conclusion

Our study inferred that the clinical symptoms persist in participants until 6 months and a multi-system involvement is seen in the post-COVID period. Thus, the findings necessitate long-term, regular follow-ups. Since the post covid sequel of covid-19 is not explicit, it is important to continue studying the long-term effects of Covid-19 on the body to better understand how to treat and prevent Post Covid syndrome. In conclusion, our study suggests a comprehensive healthcare approach for the all-inclusive management of patients.

## Statement and Declaration

### Declaration on Competing Interest (If present, give more details)

The authors declare no competing interest.

### Funding

The research was supported by the Indian Council of Medical Research, Department of Health Research, Ministry of Health and Family Welfare, Government of India. Specifically, the funding was provided through grant No.CTU/Cohort study/17/1025/2021/ECD. We are grateful for their support in helping us advance our study and improve our understanding of the important issues at hand.

## Acknowledgements

The authors would like to thank Dean, Maulana Azad Medical College for providing support in the course of the conduction of the study. We also express thanks to the team of Doctors For You, Radiology Department Lok Nayak Hospital and NDTB, Delhi for providing necessary diagnostic support.

## Ethics Statement

The Institutional Ethics Committee, Maulana Azad Medical College and Associated Hospitals, New Delhi issued approval vide letter no F.1/IEC/MAMC/87/05/2021/No526). All the eligible study participants were informed about the purpose, risks and benefits of participating in the study. They were assured of complete confidentiality and were explained the option of withdrawing from the study at any point in time if they desire to do so. Written and informed consent was obtained from all the study participants. Participants were referred to Lok Nayak Hospital for medical/teleconsultation and follow-ups.

## Consent for publication

Not Applicable

## Data Availability Statement

The data that support the findings of this study are available on request from the corresponding author. The data are not publicly available due to privacy or ethical restrictions.

## Authors contribution

All the authors contributed to the development of the Concept, literature search and design of the study. Nidhi Bhatnagar, MM Singh, Hitakshi Sharma, Gurmeet Singh, Suruchi Mishra, Shivani Rao and Amod Borle contributed to the data acquisition and analysis of the study. Suruchi Mishra, Hitakshi Sharma and Nidhi Bhatnagar prepared the manuscript. Nidhi Bhatnagar contributed to manuscript editing and MM Singh in review.

## Notes

### Competing Interest Statement

The authors have declared no competing interest.

## REFERENCES

1. WHO Situation Reports. Available from: https://www.who.int/docs/default-source/coronaviruse/situation-reports/20200423-sitrep-94-covid-19.pdf. Last accessed on 4th Aug, 2023.

2. WHO Coronavirus tracker. Available from: https://covid19.who.int/. Last accessed on 4th Aug, 2023.

3. Interim Guidance on Ending Isolation and Precautions for Adults with COVID-19. Available from: https://www.cdc.gov/coronavirus/2019-ncov/hcp/duration-isolation.html. Last accessed on 4th Aug, 2023.

4. Sisó-Almirall, A., Brito-Zerón, P., Ferrín, L. C., Kostov, B., Moreno, A. M., Mestres, J., et al. (2021). Long Covid-19: Proposed Primary Care Clinical Guidelines for Diagnosis and Disease Management. International Journal of Environmental Research and Public Health, 18(8).

5. NICE guideline on long COVID -The Lancet Respiratory Medicine [Internet]. Available from: https://www.thelancet.com/article/S2213-2600(21)00031-X/fulltext. Last accessed on 4th Aug, 2023.

6. Page, M. The British Medical Association Illustrated Medical Dictionary, 4th ed.; Dorling Kindersley: London, UK, 2018; pp. 177–536.

7. Al-Jahdhami I, Al-Naamani K, Al-Mawali A. The Post-acute COVID-19 Syndrome (Long COVID). Oman Med J. 2021 Jan 26;36(1):e220.

8. Havervall S, Rosell A, Phillipson M, Mangsbo SM, Nilsson P, Hober S, et al. Symptoms and Functional Impairment Assessed 8 Months After Mild COVID-19 Among Health Care Workers. JAMA. 2021 May 18;325(19):2015–6.

9. Post Covid Conditions: Overview for healthcare providers. Available from: https://www.cdc.gov/coronavirus/2019-ncov/hcp/clinical-care/post-covid-conditions.html. Last accessed on 4th Aug, 2023.

10. Naik S, Haldar SN, Soneja M, Mundadan NG, Garg P, Mittal A, et al. Post COVID-19 sequelae: A prospective observational study from Northern India. Drug Discov Ther. 2021 Nov 21;15(5):254–260. doi: 10.5582/ddt.2021.01093.

11. WHO case report form. Available from: https://www.who.int/publications/i/item/global-covid-19-clinical-platform-case-report-form-(crf)-for-post-covid-conditions-(post-covid-19-crf-). Last accessed on 4th Aug, 2023.

12. Huang, L., Yao, Q., Gu, X., Wang, Q., Ren, L., Wang, Y., et al (2021). 1-year outcomes in hospital survivors with COVID-19: A longitudinal cohort study. The Lancet, 398(10302), 747–758.

13. Selvakumar J, Havdal LB, Drevvatne M, Brodwall EM, Lund Berven L, Stiansen-Sonerud T, et al. Prevalence and Characteristics Associated with Post–COVID-19 Condition Among Nonhospitalized Adolescents and Young Adults. JAMA Network Open. 2023 Mar 30;6(3):e235763.

14. Fischer A, Zhang L, Elbéji A, Wilmes P, Oustric P, Staub T, et al. Long COVID Symptomatology After 12 Months and Its Impact on Quality of Life According to Initial Coronavirus Disease 2019 Disease Severity. Open Forum Infect Dis. 2022 Aug 5;9(8):ofac397. doi: 10.1093/ofid/ofac397.

15. Arjun, M. C., Singh, A. K., Roy, P., Ravichandran, M., Mandal, S., Pal, D., Das, K., et al. (2023). Long COVID following Omicron wave in Eastern India—A retrospective cohort study. Journal of Medical Virology, 95(1). 10.1002/jmv.28214

16. Menges, D., Ballouz, T., Anagnostopoulos, A., Aschmann, H. E., Domenghino, A., Fehr, J. S., et al. (2021). Burden of post-COVID-19 syndrome and implications for healthcare service planning: A population-based cohort study. PLOS ONE, 16(7), e0254523. 10.1371/journal.pone.0254523

17. Cioboata, R., Nicolosu, D., Streba, C. T., Vasile, C. M., Olteanu, M., Nemes, A., et al. (2022). Post-COVID-19 Syndrome Based on Disease Form and Associated Comorbidities. Diagnostics, 12(10). 10.3390/diagnostics12102502.

18. Alfadda, A. A., Rafiullah, M., Alkhowaiter, M., Alotaibi, N., Alzahrani, M., Binkhamis, K., et al. (2022). Clinical and biochemical characteristics of people experiencing post-coronavirus disease 2019-related symptoms: A prospective follow-up investigation. Frontiers in Medicine, 9, 1067082. 10.3389/fmed.2022.1067082.

19. Gameil MA, Marzouk RE, Elsebaie AH, Rozaik SE. Long-term clinical and biochemical residue after COVID-19 recovery. Egypt Liver J. 2021;11(1):74. doi: 10.1186/s43066-021-00144-1. Epub 2021 Sep 12.

20. Ruenjaiman, V., Sodsai, P., Kueanjinda, P., Bunrasmee, W., Klinchanhom, S., Reantragoon, R., et al. (2022). Impact of SARS-CoV-2 infection on the profiles and responses of innate immune cells after recovery. Journal of Microbiology, Immunology and Infection, 55(6), 993–1004. 10.1016/j.jmii.2022.09.001

21. Fogante, M., Cavagna, E., Rinaldi, G. (2022). COVID-19 follow-up: Chest X-ray findings with clinical and radiological relationship three months after recovery. Radiography (London, England : 1995), 28(2), 531–536. 10.1016/j.radi.2021.10.012

22. Øvrebotten, T., Myhre, P. L., Grimsmo, J., Mecinaj, A., Trebinjac, D., Nossen, M. B., et al. (2022). Changes in cardiac structure and function from 3 to 12 months after hospitalization for COVID-19. Clinical Cardiology, 45(10), 1044–1052. 10.1002/clc.23891

