## Supplemental Tables for "Prevalence and pattern of Post Covid-19 symptoms in recovered patients of Delhi: A Population-Based study"

Annexure I: Distribution of study participants according to biochemical investigations.

| Biochemical Investigations | Baseline (N=351) | Follow up 1 (N=281) | | Follow up 2 (N=187) | | |
| --- | --- | --- | --- | --- | --- | --- |
|  | Raised HbA1c | Prevalence | Incidence | Prevalence | Incidence | |
| Raised HbA1c  (> 6.5%) | 23 (6.5) | 20 (7.1) | 10 (3.5) | 36 (19.2) | 13 (6.9) | |
|  | Baseline (N=330) | Follow up 1(N=337) | | Follow up 2 (N=350) | | |
|  | Elevated TSH | Prevalence | Incidence | Prevalence | Incidence | |
| Raised TSH(>4.68mIU/L) | 46 (13.9) | 49 (14.5) | 31 (9.2) | 56 (16.0) | 23 (6.6) | |
| Liver Function Test | Baseline (N=362) | Follow up 1 (N=357) | | Follow up 2 (N=364) | | |
|  |  | Prevalence | Incidence | Prevalence | | Incidence |
| AST/SGOT  (>59 IU/L) | 35 (9.8) | 25 (7) | 15 (4.2) | 23 (6.4) | | 13 (3.6) |
| ALT/SGPT  (>50 IU/L) | 73 (20.4) | 70 (19.6) | 30 (8.4) | 52 (14.6) | | 17 (4.8) |
| ALP  (>126 IU/L) | 57 (16.0) | 68 (19) | 45 (12.6) | 60 (16.8) | | 25 (7.0) |
| Kidney Function Test | Baseline (N=363) | Follow up 1 (N=357) | | Follow up 2 (N=364) | | |
|  |  | Prevalence | Incidence | Prevalence | | Incidence |
| Creatinine  (>1.3mg/ dl) | 6 (1.7) | 7 (2.0) | 3 (0.8) | 6 (1.6) | | 4 (1.1) |
| Urea  (>43 mg/dl) | 12 (3.3) | 11 (3.1) | 9 (2.5) | 10 (2.7) | | 6 (1.6) |
| Uric Acid( 8.5mg/dl) | 10 (2.8) | 9 (2.5) | 7 (1.9) | 6 (1.7) | | 5 (1.4) |
| Peripheral Blood Smear | Baseline | Follow up 1 | | Follow up 2 | | |
|  |  | Prevalence | Incidence | Prevalence | | Incidence |
| Transformed Lymphocytes | 31 (8.4) | 6 (1.6) | 4 (1.1) | 4 (1.1) | | 3 (0.8) |
| Activated Monocytes | 14 (3.8) | 5 (1.4) | 4 (1.1) | 4 (1.1) | | 3 (0.8) |
| Absolute Neutrophil Count (Abnormally high) | 65 (17.6) | 81 (22.0) | 61 (16.5) | 78 (21.1) | | 43 (11.7) |
| Absolute Eosinophil Count (Abnormally high) | 32 (8.7) | 36 (9.8) | 29 (7.9) | 26 (7.0) | | 17 (4.6) |

Annexure II: Hematological and Biochemical reference value.

| Blood Profile | Reference Value |
| --- | --- |
| HbA1C | 5.7% |
| KFT | Reference Value |
| Creatinine | 0.7-1.3 mg/dl |
| Urea | 19-43 mg/dl |
| Uric Acid | 3.5-8.5 mg/dl |
| LFT | Reference Value |
| AST | 17-59 IU/L |
| ALP | 38-126 IU/L |
| ALT | 5-50 IU/L |
| TFT | Reference Value |
| TSH | 0.465-4.68 IU/L |
| T3 | 4.26-8.10 IU/L |
| T4 | 10.0-28.2 IU/L |
| P/S | Reference Value |
| ANC | 2500-7000 |
| AEC | 0-500 |
